# Real-world safety and effectiveness of olaparib maintenance after first-relapse platinum-sensitive ovarian cancer in Japan: a multicenter historical cohort (JGOG3026)

**DOI:** 10.1101/2025.09.06.25335250

**Authors:** Kosuke Yoshihara, Terumi Tanigawa, Shin Nishio, Hiroyuki Kanao, Yasunori Yoshino, Ayako Mochizuki, Sari Nakao, Tatsuo Kagimura, Hiroshi Tanabe, Hiroaki Kajiyama, Muneaki Shimada, Aikou Okamoto, Takayuki Enomoto

## Abstract

**Objective:** The primary objective of the study was to evaluate the safety and effectiveness of olaparib maintenance therapy in Japanese patients with platinum-sensitive first-relapsed ovarian cancer. The secondary objective was to survey the treatment given after the olaparib maintenance therapy.

**Methods:** The JGOG3026 study was a nationwide, multicenter historical cohort study across 43 sites. Patients receiving olaparib maintenance therapy after platinum-based chemotherapy between Jan 2018 and Jul 2020 were included as modified intention-to-treat (mITT1). Those who received any post-olaparib therapy were extracted as mITT2. The primary endpoint was adverse events; secondary endpoints included progression-free survival (PFS), the time to first subsequent therapy (TFST), progression-free survival to the second progression (PFS2), and overall survival (OS).

**Results:** In the mITT1 (n = 413), any adverse event occurred in 41.6%; anemia was the most common (28.8%). MDS occurred in 1.0% and no AML was observed during a mean follow-up of 34.1 months. The 24-month PFS and OS rates were 31.7% and 70.1%, respectively. Median PFS and OS were 12 months (95% confidence interval [CI]: 9.9-13) and 39 months (95% CI, 33–45). The 24-month TFST and PFS2 rates were 33.3% and 34.3%. In the mITT2 (n=284), chemotherapy after olaparib therapy was used in 97.2%, with an overall response rate of 22.5%.

**Conclusion:** Olaparib maintenance therapy showed acceptable safety and effectiveness in Japanese clinical practice. Monitoring for secondary hematologic malignancies is needed after the start of olaparib maintenance therapy.

**Synopsis:** Our multicenter collaborative historical cohort study confirmed acceptable safety and effectiveness of olaparib maintenance therapy in Japanese patients with platinum-sensitive first-relapsed ovarian cancer; the incidence of AML/MDS was comparable to reports from Western cohorts.

## INTRODUCTION

In January 2018, olaparib, a poly (ADP-ribose) polymerase (PARP) inhibitor, was approved for the first time in Japan for the maintenance therapy for relapsed ovarian cancer sensitive to platinum-containing anti-cancer drug (platinum-sensitive) regardless of *BRCA*1/2 mutation status. Although evidence of the safety and efficacy of olaparib in patients with platinum-sensitive relapsed ovarian cancer is based on the Study 19 [1–3] and SOLO2 [4–6] studies, the number of Japanese participants in these studies is 0 and 8, respectively. The long-term incidence of adverse events, such as secondary malignancies, had not been clarified in Japan. Therefore, the evaluation of the safety and effectiveness of olaparib maintenance therapy in Japan could be an urgent issue.

When disease progression is noted during olaparib maintenance therapy, the olaparib maintenance therapy will be terminated. Chemotherapy may be considered as a subsequent treatment in patients with a favorable performance status. However, because there was no definitive evidence that platinum-based combination chemotherapy should be administered for recurrent ovarian cancer after PARP inhibitors maintenance therapy, treatment might be selected in consideration of platinum-free interval (PFI), patient’s medical history, and other data. Still, it was unclear whether the duration of olaparib treatment could be included in the PFI, either. Some mechanisms had been suggested for acquired resistance to PARP inhibitors [7], and homologous recombination repair restoration was cited as one of the examples [8]. Because homologous recombination repair deficiency was a common feature of platinum-sensitive ovarian cancer, platinum-containing compounds might not be highly effective against tumors with acquired resistance to PARP inhibitors either.

Motivated by the limited participation of Japanese patients in pivotal trials and uncertainties regarding the effectiveness of post-PARP chemotherapy, we initiated this study in 2020 to assess the real-world safety and efficacy of olaparib maintenance therapy in Japan [9], as well as to conduct a fact-finding survey on the subsequent treatments administered.

## MATERIALS AND METHODS

### Patients and study design

A multicenter collaborative historical cohort study (JGOG3026; UMIN 000041422) was conducted in Japan between August 2020 and August 2024. We included patients who initiated olaparib maintenance therapy after platinum-based combination chemotherapy for platinum-sensitive first-relapsed ovarian, fallopian tube, or primary peritoneal cancer between January 2018 and July 2020. Participants were enrolled for 6 months from January 2021 in 43 Japanese Gynecologic Oncology (JGOG) institutions throughout Japan (Table 1), and the follow-up period was 26 months after the end of enrollment (August 2023). Clinical outcomes were updated prospectively every 12 months in the electronic data capture system.

**Table 1.** Clinicopathological characteristics of subjects in JGOG3026.

|  | **mITT1 (n = 413)** | **mITT2 (n = 284)** |
| --- | --- | --- |
| Age | 63.9 ± 10.5 | 63.8 ± 10.8 |
| History of Cancer | 57 (13.8%) | 35 (12.3%) |
| gBRCA test | 44 (10.7%) | 27 (9.5%) |
| Primary tumor location |  |  |
| Ovary | 312 (75.5%) | 218 (76.8%) |
| Fallopian tubes | 48 (11.6%) | 33 (11.6%) |
| Primary peritoneum | 47 (11.4%) | 29 (10.2%) |
| Other | 6 (1.5%) | 4 (1.4%) |
| Histology type |  |  |
| Serous | 338 (81.8%) | 232 (81.7%) |
| Clear Cell | 19 (4.6%) | 15 (5.3%) |
| Endometrioid | 26 (6.3%) | 15 (5.3%) |
| Mucinous | 1 (0.2%) | 1 (0.4%) |
| Others | 29 (7.0%) | 21 (7.4%) |
| Stage before recurrence |  |  |
| I | 29 (7.0%) | 18 (6.3%) |
| II | 32 (7.8%) | 18 (6.3%) |
| III | 247 (59.8%) | 180 (63.4%) |
| IV | 102 (24.7%) | 68 (23.9%) |
| Unknown | 3 (0.7%) | 0 (0.0%) |
| Response to previous platinum therapy |  |  |
| Complete response (CR) | 118 (28.6%) | 73 (25.7%) |
| Partial response (PR) | 293 (70.9%) | 210 (73.9%) |
| Not evaluable (NE) | 2 (0.5%) | 1 (0.4%) |
| Chemotherapy regimen |  |  |
| Paclitaxel + Carboplatin | 227 (55.0%) | 151 (53.2%) |
| dose-dense Paclitaxel + Carboplatin | 19 (4.6%) | 12 (4.2%) |
| Pegylated Liposomal Doxorubicin + Carboplatin | 71 (17.2%) | 53 (18.7%) |
| Gemcitabine + Carboplatin | 33 (8.0%) | 28 (9.9%) |
| Others | 63 (15.2%) | 40 (14.0%) |
| Bevacizumab use | 149 (36.1%) | 113 (39.8%) |
| Platinum-free interval |  |  |
| < 12 months | 133 (33.8%) | 110 (40.6%) |
| ≥ 12 months | 260 (66.2%) | 161 (59.4%) |

The target sample size was set to 300 based on the number of patients who can be enrolled under actual clinical practice in Japan. Treatment selection was basically performed according to the Japan Society of Gynecologic Oncology guidelines for the treatment of ovarian cancer, fallopian tube cancer, and primary peritoneal cancer [10]. All evaluations were scheduled according to the clinical practice standards of each institution. We collected data including patient characteristics, clinicopathological data, clinical outcomes, and adverse events. We also collected germline *BRCA* (g*BRCA*) status, but not homologous recombination deficiency (HRD) status. The stage before recurrence was determined based on the FIGO 2014 staging system [11].

The study was conducted in accordance with the Declaration of Helsinki and the Ethical Guidelines for Medical and Health Research Involving Human Subjects. After approval at Niigata University (G2020-0016), approval from the institutional review board of each participating JGOG institution was obtained before the initiation of the study. Written informed consent was obtained for participation in this study and the use of the information about the g*BRCA* status.

### Study outcome

The primary endpoint was the incidence of adverse events in patients who received olaparib maintenance therapy for platinum-sensitive first-relapsed ovarian cancer. The secondary endpoints were progression-free survival (PFS), time to first subsequent therapy (TFST), progression-free survival to the second progression (PFS2), and overall survival (OS) in patients who received olaparib maintenance therapy for platinum-sensitive first-relapsed ovarian cancer [4]. In addition, the details and effects of treatments for recurrence after olaparib maintenance therapy for platinum-sensitive first-relapsed ovarian cancer were assessed.

### Study population

The inclusion criteria were as follows: Patients with first-relapsed ovarian cancer (including fallopian tube cancer and primary peritoneal cancer) who received platinum-based combination chemotherapy and then started to undergo olaparib maintenance therapy between January 2018 and July 2020; Patients aged 20 years or older (without upper age limit) at the time of enrollment. Patients were excluded based on the following criteria: Patients with platinum-resistant relapsed ovarian cancer; Chemotherapy-treated patients who have not achieved a complete response (CR) or partial response (PR) since relapse; Patients who were previously treated with poly (ADP-ribose) polymerase (PARP) inhibitors before olaparib maintenance therapy; Patient receiving maintenance therapy with olaparib and bevacizumab; Patients disqualified from participation in the study by the investigator. Patients who started to receive olaparib maintenance therapy between January 2018 and July 2020, following platinum-based combination chemotherapy for the treatment of first-relapsed ovarian cancer, were assigned as the modified intention-to-treat (mITT1) group (Figure 1). In addition, patients who received any treatment for recurrence after olaparib maintenance were extracted as mITT2.

**Figure 1.**
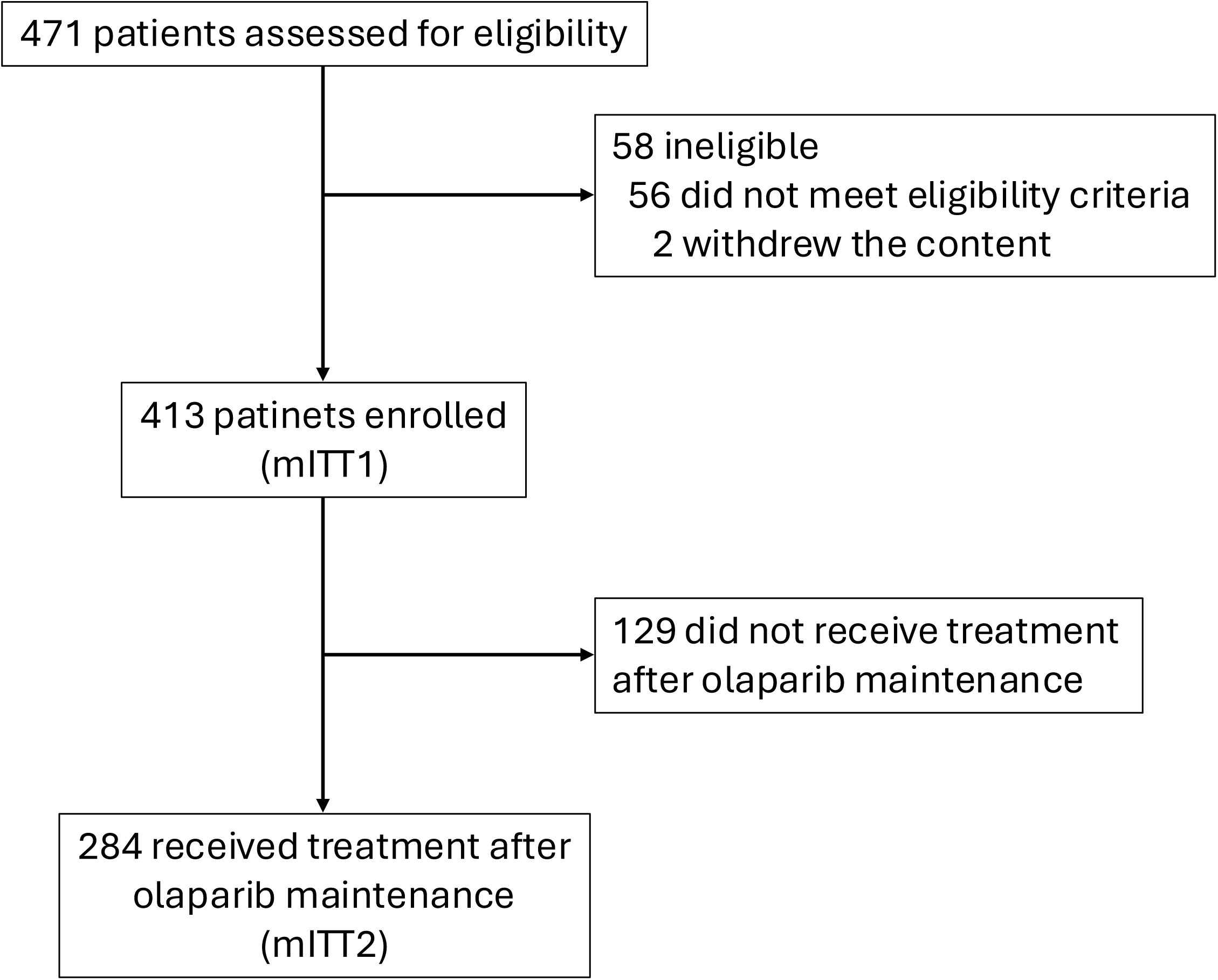
Flow chart of JGOG3026 study

### Definition of adverse events

An adverse event was defined as any unfavorable medical event that occurred during or after the clinical study. According to AstraZeneca’s drug risk management plan of olaparib, myelosuppression (including anemia [32.4%], neutropenia [16.8%], leukopenia [13.8%], thrombocytopenia [9.5%], and lymphopenia [4.0%]) and interstitial lung disease (0.8%) were considered significant identified risks during clinical trials, and secondary malignant tumors, embryotoxicity, and patients with renal impairment were cited as critical potential risks. Therefore, safety information about CTCAE Grade ≥ 3 adverse events and secondary malignant tumors [myelodysplastic syndrome (MDS), acute myeloid leukemia (AML), and others] with unknown incidence, which were classified as critical potential risks, was collected mainly during the study.

### Statistical analysis

The data were analyzed using SAS version 9.4 (SAS Institute, Inc., Cary, NC, USA) and R software v.4.4.3 (R Foundation, Vienna, Austria). The frequency of adverse events was calculated as a percentage, along with a 95% confidence interval (CI). The cumulative survival rates for PFS, TFST, PFS2, and OS were estimated by the Kaplan–Meier method. In stratified analysis, Cox proportional hazards models were used to estimate hazard ratios and their 95% confidence intervals. PFS was defined as the time from the start date of olaparib maintenance therapy to disease progression confirmed or death from any cause, whichever occurs first. TFST was defined as the time from the start date of olaparib maintenance therapy to initiation of first subsequent therapy or death, whichever occurred first. PFS2 was defined as the time from the start date of olaparib maintenance therapy to second objective progression assessed by RECIST v1.1 or death, whichever occurred first. Patients who discontinued the study without having an event were analyzed as censored cases at the time of discontinuation. Imputation of missing values was not performed for the secondary endpoints. Data are expressed as mean (SD) for continuous variables, and frequencies and percentages for discrete variables, unless specifically mentioned, and 95% confidence intervals were calculated by the Clopper-Pearson method.

## RESULTS

### Patient characteristics

The study was conducted from August 2020 to August 2024. The enrollment period was initially planned for 4 months but was extended to 6 months to reach the required sample size. A total of 471 patients were enrolled, but 58 patients were excluded from the intention-to-treat (ITT) set according to the eligibility criteria. A total of 413 patients were included in the mITT1 set (Figure 1). To assess the details and effectiveness of treatment for recurrence after olaparib maintenance therapy, 284 patients who received any treatment for recurrence after olaparib maintenance were extracted as the mITT2 set. The patient characteristics of mITT1 and mITT2 are shown in Table 1. Patients in the mITT1 set had a mean age of 63.9 years. Of the 413 patients, 14.8% had stage I/II disease, and 84.5% had stage III/IV disease before recurrence. The mean time from the start of olaparib treatment to the last follow-up was 34.1 ± 15.4 months in the mITT1.

### Primary endpoint

The incidence of adverse events in Japanese patients who received olaparib maintenance therapy for platinum-sensitive first-relapsed ovarian cancer is shown in Table 2. Of 413 patients, 172 (41.6%) had CTCAE Grade ≥ 3 adverse events during olaparib maintenance therapy, and anemia (28.8%; [95%CI 24.5-33.4%]) was the most common adverse event in mITT1. One patient (0.2%) developed interstitial lung disease in mITT1. As secondary malignancies, MDS was diagnosed in 4 patients (1.0%) of mITT1, but AML or other solid malignant tumors were not detected in mITT1 during the average follow-up period of 34.1 months. The frequencies of dose reductions and interruptions owing to adverse events were 13% and 39% in mITT1, respectively (Table S2).

**Table 2.** Incidence of adverse events in patients who received olaparib maintenance therapy for platinum-sensitive first-relapsed ovarian cancer.

|  | **mITT1**<br>**(n = 413)** |  | **mITT2**<br>**(n = 284)** |  |
| --- | --- | --- | --- | --- |
|  | N (%) | 95%CI | N (%) | 95%CI |
| Anemia | 119 (28.8) | 24.5-33.4 | 81 (28.5) | 23.3-34.2 |
| White blood cell decreased | 11 (2.7) | 1.3-4.7 | 10 (3.5) | 1.7-6.4 |
| Neutrophil count decreased | 36 (8.7) | 6.2-11.9 | 24 (8.5) | 5.5-12.3 |
| Febrile neutropenia | 0 (0) | 0.0-0.9 | 0 (0) | 0.0-1.3 |
| Lymphocyte count decreased | 1 (0.2) | 0.0-1.3 | 0 (0) | 0.0-1.3 |
| Platelet cell decreased | 16 (3.9) | 2.2-6.2 | 12 (4.2) | 2.2-7.3 |
| Hypertension | 0 (0) | 0.0-0.9 | 0 (0) | 0.0-1.3 |
| Interstitial lung disease | 1 (0.2) | 0.0-1.3 | 0 (0) | 0.0-1.3 |
| MDS | 4 (1.0) | 0.3-2.5 | 2 (0.7) | 0.1-2.5 |
| AML | 0 (0.0) | 0.0-0.9 | 0 (0) | 0.0-1.3 |
| Others | 35 (8.5) | 6.0-11.6 | 19 (6.7) | 1.7-6.4 |
| Total adverse events | 172 (41.6) | 36.8-46.6 | 114 (40.1) | 34.4-46.1 |

### Secondary endpoints

We performed Kaplan–Meier survival analyses for PFS, TFST, and OS in the mITT1 set (Figures 2 and 3A). The Kaplan-Meier survival curve demonstrated that the 12-month and 24-month PFS rates were 48.5% (95% CI: 43.5-53.3) and 31.7% (95% CI: 27.2-36.3), respectively (Figure 2A). The 12-month OS and 24-month OS were also 92.9% (95%CI, 90.0-95.0) and 70.1% (95%CI, 65.4-74.3%) (Figure 2B). The median survival times for PFS, TFST, and OS were 12 months (95% CI: 9.9-13.0), 13 months (95% CI: 12.0-15.0), and 39 months (95% CI: 33.0-45.0), respectively. We also examined PFS2 in the mITT2 set (Figure 3B). The median survival of PFS2 was 34.3% (95% CI: 28.8-39.9).

**Figure 2.**
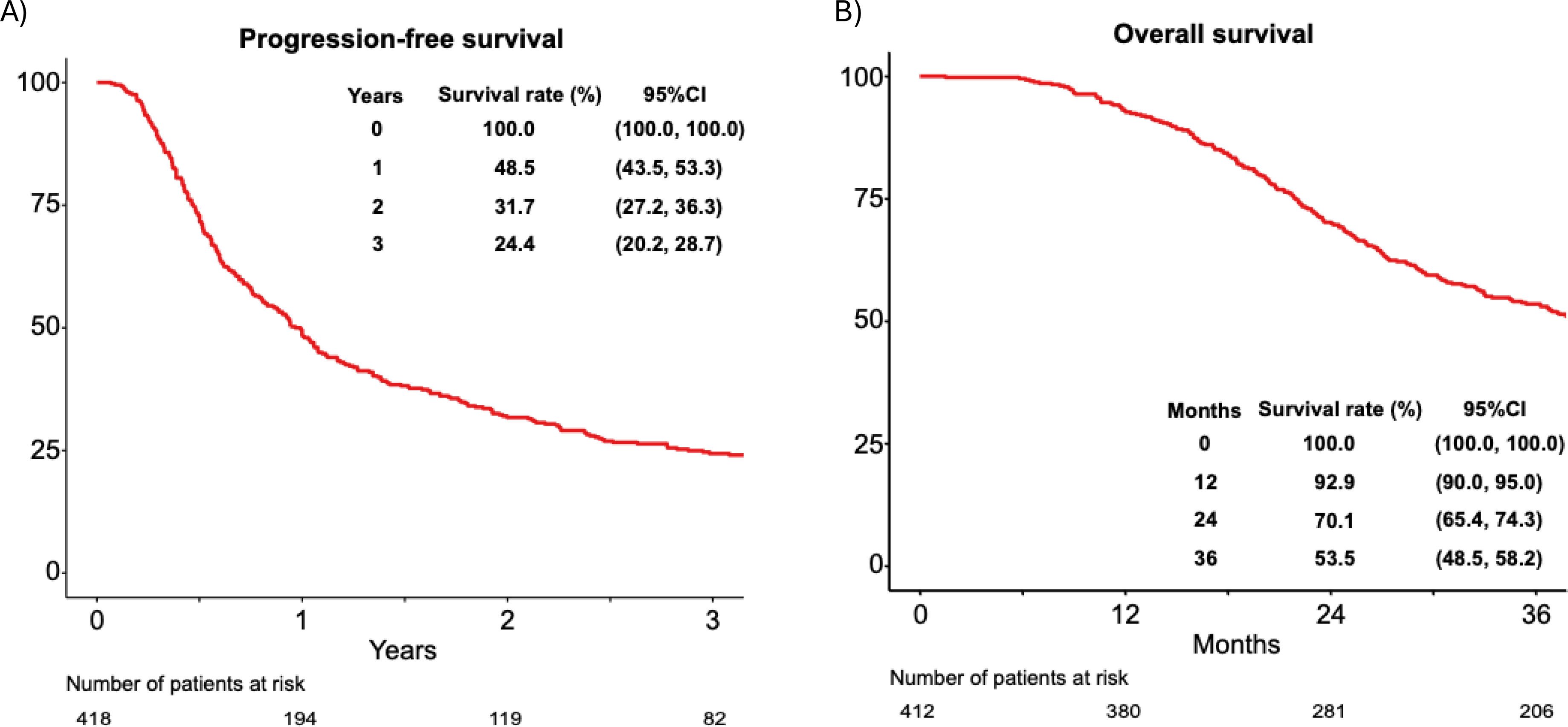
Kaplan-Meier curves for (A) progression-free survival and (B) overall survival in the mITT1 cohort. Tick marks denote censoring. Numbers at risk are shown below each panel.

**Figure 3.**
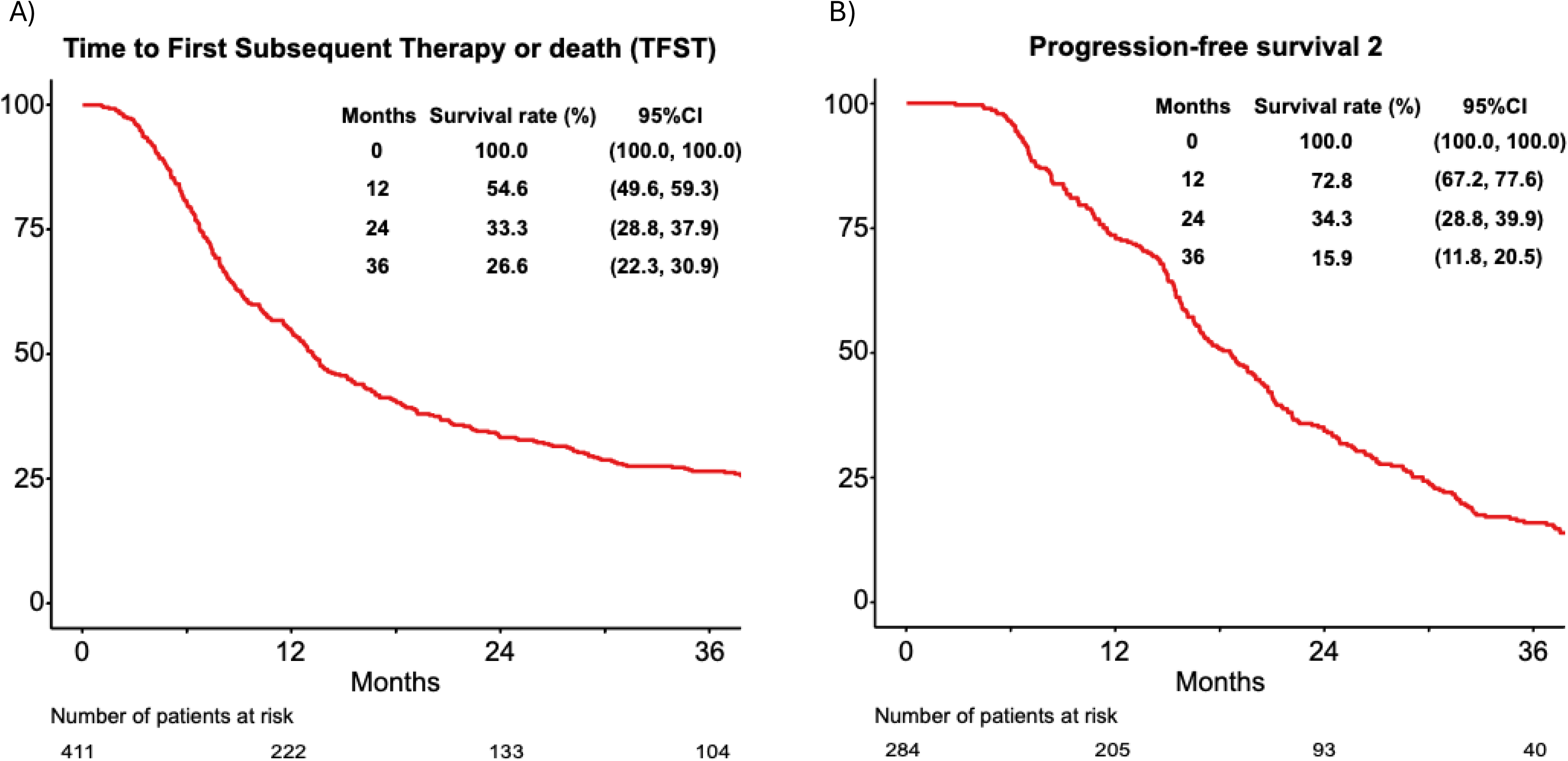
Kaplan-Meier curves for (A) time to first subsequent therapy or death (TFST) in the mITT1 cohort and (B) progression-free survival to the second progression (PFS2) in the mITT2 cohort.

Next, we stratified the patients based on chemotherapy response, germline *BRCA1/2 (gBRCA)* status, histology, and FIGO stage. According to the response assessment, 405 patients were divided into the two subgroups: complete response (CR) group (n = 117) and partial response (PR) group (n = 289) (Figure S1A). The CR group had longer PFS than the PR group (HR 0.68 [95% CI, 0.53-0.87]). Although the results of germline *BRCA* testing were available in only 43 patients, PFS was significantly longer in the patients with g*BRCA* pathogenic variants compared to those with g*BRCA* wildtype (HR 0.35 [95% CI, 0.16-0.75]) (Figure S1B). High-grade serous histologic type was marginally associated with PFS compared to other histologic types (HR 0.77; 95% CI, 0.59-1.00). On the other hand, the FIGO stage before recurrence was not associated with PFS (Figure S2). In the mITT1 population, 29 patients received secondary debulking surgery (SDS). The SDS group showed longer PFS compared to the non-SDS group (HR 0.28 [95% CI, 0.16-0.50]) (Figure S3). The association between each factor and OS was listed in Table S3. Chemotherapy response, g*BRCA* status, histology, and SDS were also associated with OS.

Finally, we investigated the details and effectiveness of subsequent treatment after olaparib maintenance therapy for platinum-sensitive first-relapsed ovarian cancer. In the mITT2 population, 97.2% of patients received chemotherapy (Table 3), and the response rate was 22.5%. The non-platinum regimen (n=162) was frequently selected compared to the platinum regimen (n = 78) for recurrent ovarian cancer after olaparib maintenance therapy (Table S4). The response rates of the platinum or non-platinum regimens were 19.2% and 21.6%, respectively.

**Table 3.** Details of treatment for recurrence after olaparib maintenance therapy.

|  | **mITT2 (n = 284)** |
| --- | --- |
| Surgery | 16 (5.6%) |
| Chemotherapy | 276 (97.2%) |
| Regimen |  |
| Paclitaxel + Carboplatin | 65 (22.9%) |
| Pegylated Liposomal Doxorubicin + Carboplatin | 60 (21.1%) |
| Gemcitabine + Carboplatin | 33 (11.6%) |
| Others | 117 (41.2%) |
| Bevacizumab use | 123 (43.3%) |
| Assessment of chemotherapy |  |
| CR | 17 (6.0%) |
| PR | 47 (16.5%) |
| SD | 54 (19.0%) |
| non-CR/non-PD | 1 (0.4%) |
| NE | 1 (0.4%) |
| PD | 164 (57.7%) |

## DISCUSSION

In this multicenter collaborative historical cohort study, we evaluated the safety and effectiveness of olaparib maintenance therapy for platinum-sensitive first-relapsed ovarian cancer in Japan.

A significant safety concern with olaparib maintenance therapy is the occurrence of MDS/AML. In the SOLO2 study, 1 MDS (1%) and 2 AML (1%) events occurred in the olaparib arm, while 3 MDS (3%) and 1 AML (1%) events occurred with placebo [4]. The frequency of MDS/AML after olaparib maintenance therapy in our cohort was similar to that reported in Western populations. The L-MOCA study [12], which evaluated the safety and effectiveness of olaparib maintenance therapy in Asian patients (91.5% Chinese) with platinum-sensitive relapsed ovarian cancer, also reported 3 MDS/AML (1.3%) events. Population-based pharmacokinetic analyses indicate no clinically meaningful effect of race/ethnicity on olaparib treatment (https://www.ema.europa.eu/en/documents/product-information/lynparza-epar-product-information_en.pdf). The causality of MDS/AML is difficult to ascertain because all affected patients had previously been treated with a platinum regimen.

In the SOLO2, CTCAE grade 3 or higher adverse events occurred in 69 patients (36%), with anemia being the most common (19%). Dose interruptions and reductions occurred in 39% and 13% of patients in our dataset, compared with 45% and 25% in SOLO2, respectively. Overall, no major differences in adverse event profiles or dose modification rates were observed between SOLO2 and our study.

Regarding the effectiveness of olaparib maintenance therapy, we evaluated 12-month and 24-month PFS. In the SOLO2 study [4], 12-month and 24-month PFS rates were 65% (95% CI, 57.8-71.4) and 43% (35.5-50.4), respectively. Because the SOLO2 enrolled platinum-sensitive relapsed ovarian cancer patients with a *BRCA1/2* mutation, 12-month and 24-month PFS were longer in the SOLO2 than in our overall mITT1 cohort (Figure 2). When we focused on the subsets with g*BRCA* pathogenic variants, 12-month and 24-month PFS were longer in our study (Figure S1) than in the SOLO2. In the L-MOCA study [12], 6-month and 12-month PFS were 76.0% (95% CI, 69.8-81.2) and 57.1% (95% CI, 50.2-63.5). Although histology, *BRCA* mutation status, homologous recombination repair mutation status, and last line of chemotherapy differed between the L-MOCA study and our cohort, Kaplan-Meier survival curves suggested comparable PFS in the *BRCA-*mutated subset (Figure S1B). On the other hand, the OPINION study [13, 14], which evaluated olaparib maintenance therapy for platinum-sensitive relapsed ovarian cancer without g*BRCA* pathogenic variants, reported a 12-month PFS of 38.5% (95%CI, 32.7-44.3), comparable to our findings (Figure S1B). These results support broadly similar safety and effectiveness profiles relative to non-Japanese cohorts, while acknowledging differences in *BRCA* status, histology, and prior therapies. However, we must keep in mind that olaparib shows the most substantial effect against *BRCA*-mutated ovarian cancer in the previous reports and ours [4, 12].

Notably, clear-cell carcinoma is relatively more prevalent in Japan; outcomes in non-high-grade serous subsets should therefore be interpreted with caution (Figure S2A). Although SDS was also associated with better PFS and OS in our cohort (Figure S3 and Table S3), the role of secondary cytoreductive surgery remains controversial: DESKTOP III [15] demonstrated an overall-survival benefit in carefully selected patients, whereas GOG-0213 [16] did not; SOC-1 [17] showed a PFS advantage. These differences underscore the importance of case selection. Interpretation of outcomes in patients who underwent SDS should also consider selection bias, as SDS decisions were non-random.

We also examined the subsequent therapy for relapsed ovarian cancer after olaparib maintenance therapy. Non-platinum regimens were frequently selected and appeared more effective than the platinum regimen in our cohort (Table S4). Frenel *et al*. conducted the post-hoc analyses of SOLO2 to evaluate the effectiveness of subsequent chemotherapy for recurrent ovarian cancer after olaparib maintenance therapy [6]. Time to second progression, which was calculated from the date of RECIST progression to the next progression/death, was significantly shorter in the olaparib group compared with placebo (6.9 vs 12.1 months; HR 2.17). The effectiveness was most evident in patients receiving platinum-based chemotherapy (7.0 vs 14.3 months; HR 2.89), while no significant difference was observed with non-platinum regimens (6.0 vs 8.3 months). Prior PARP inhibitor treatment may reduce the effectiveness of subsequent platinum-based chemotherapy, possibly due to overlapping resistance mechanisms such as drug target-related resistance, restoration of homologous recombination, and restoration of replication fork stability [18].

This study has limitations, including incomplete ascertainment of *BRCA* status and potential selection and survivor biases in analyses of post-olaparib therapies. Regimen choice after olaparib was non-random and likely influenced by clinical judgment and patient factors. While we report rates and survival estimates with appropriate censoring, longer follow-up would be needed to assess MDS/AML because a comprehensive survey of the AEs of olaparib using the Japanese Adverse Reaction Reporting (JADER) database highlighted the late-onset nature of MDS/AML [19].

In conclusion, olaparib maintenance therapy is a safe and effective treatment for Japanese patients with platinum-sensitive first-relapsed ovarian cancer.

## Data Availability

All data produced in the present study are available upon reasonable request to the authors.

## Acknowledgements

We thank all patients and their families. We also thank the staff from the Japanese Gynecologic Oncology Group (JGOG). The JGOG research fund was used to conduct this study.

