## Supplementary figures and images for "Real-world safety and effectiveness of olaparib maintenance after first-relapse platinum-sensitive ovarian cancer in Japan: a multicenter historical cohort (JGOG3026)"

### Figure S1

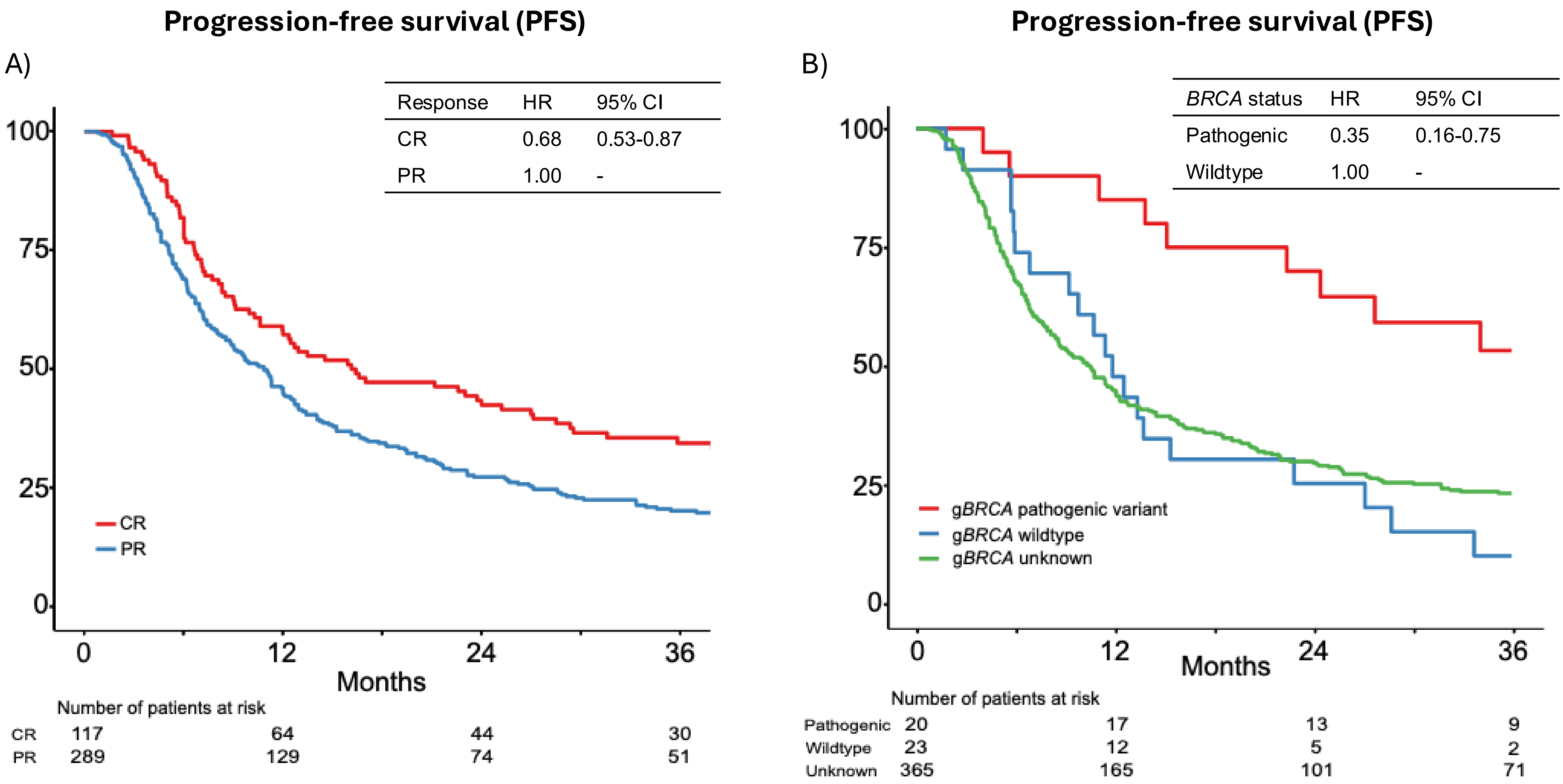

### Figure S2

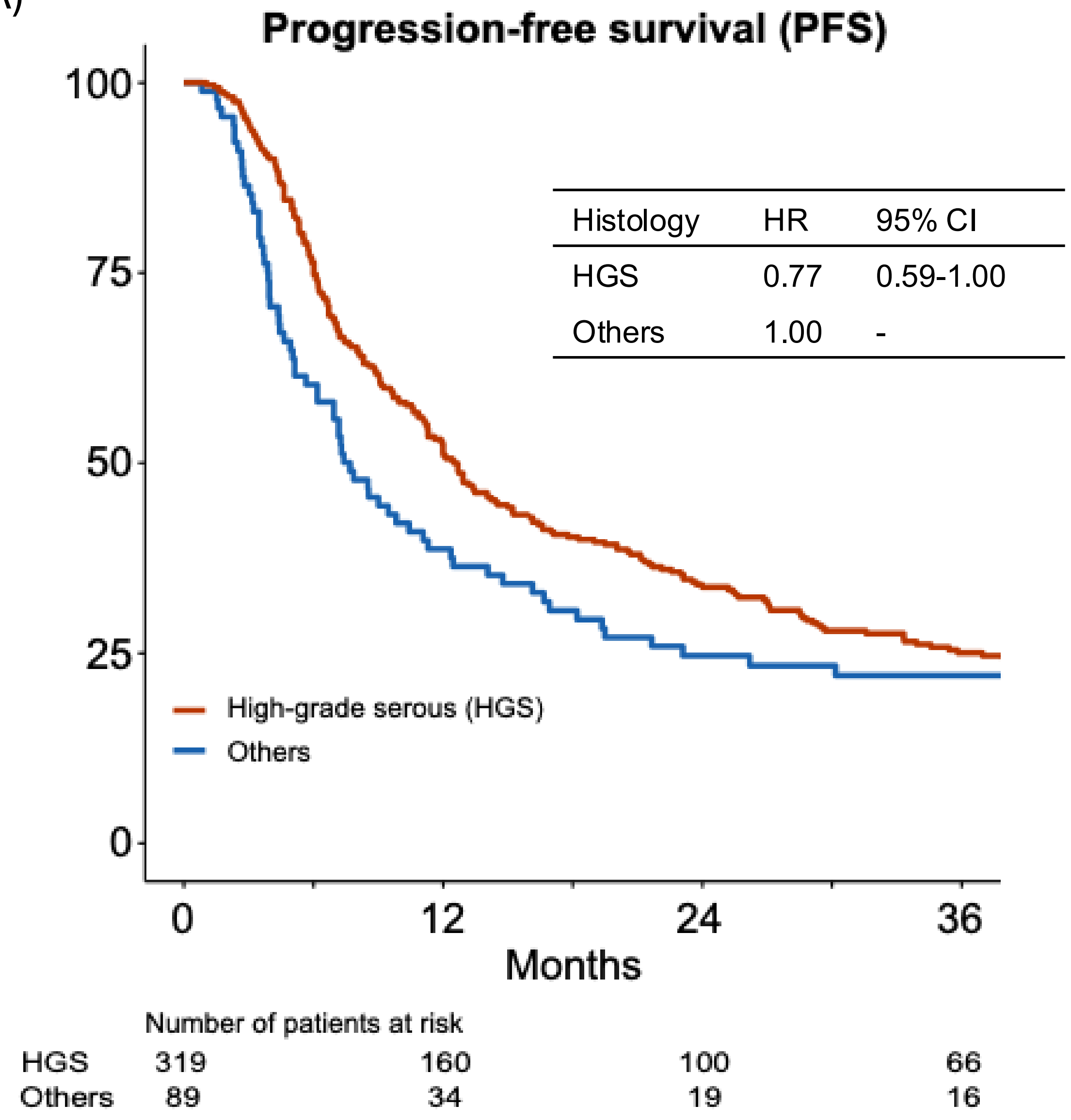

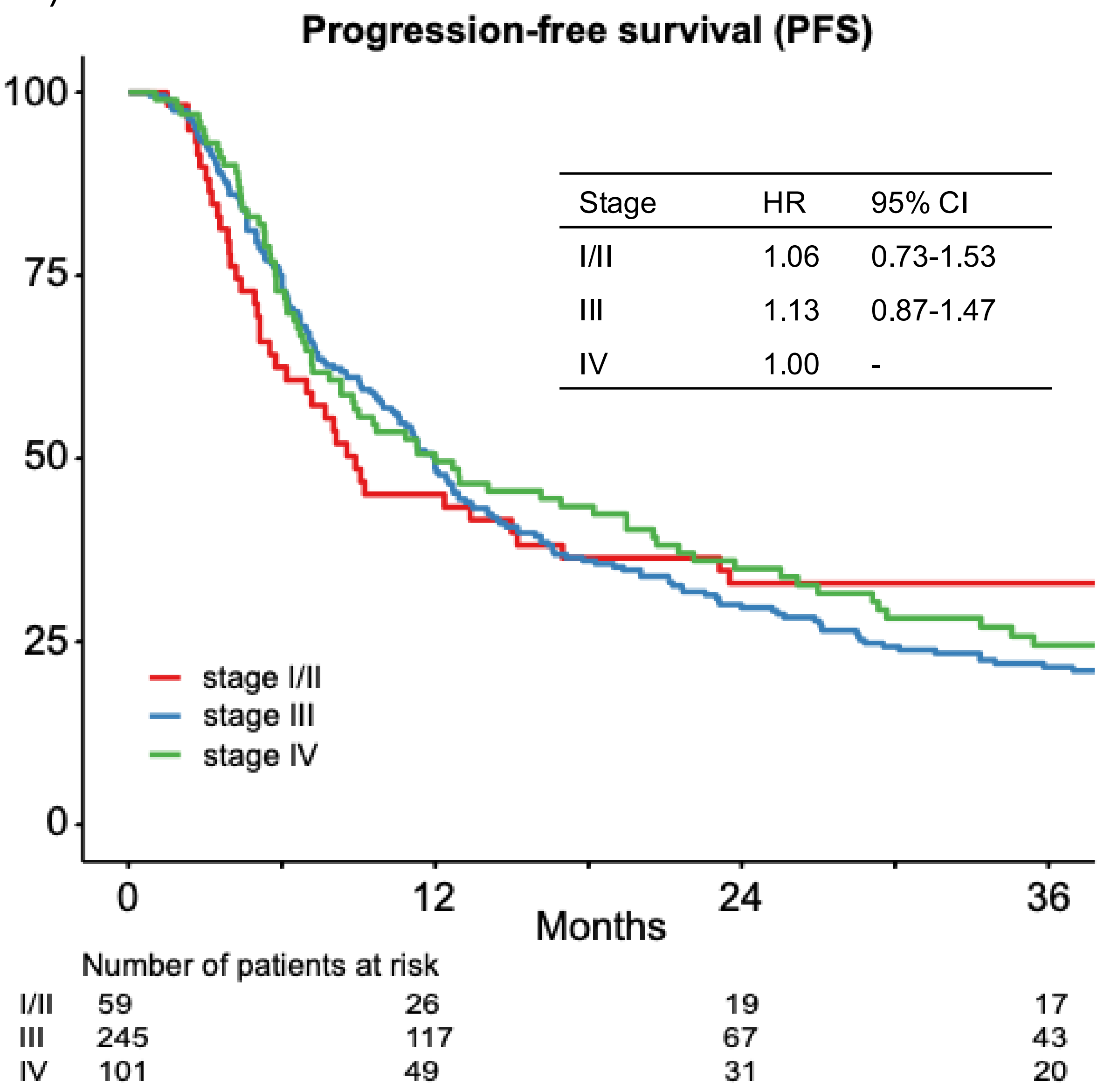

### Figure S3

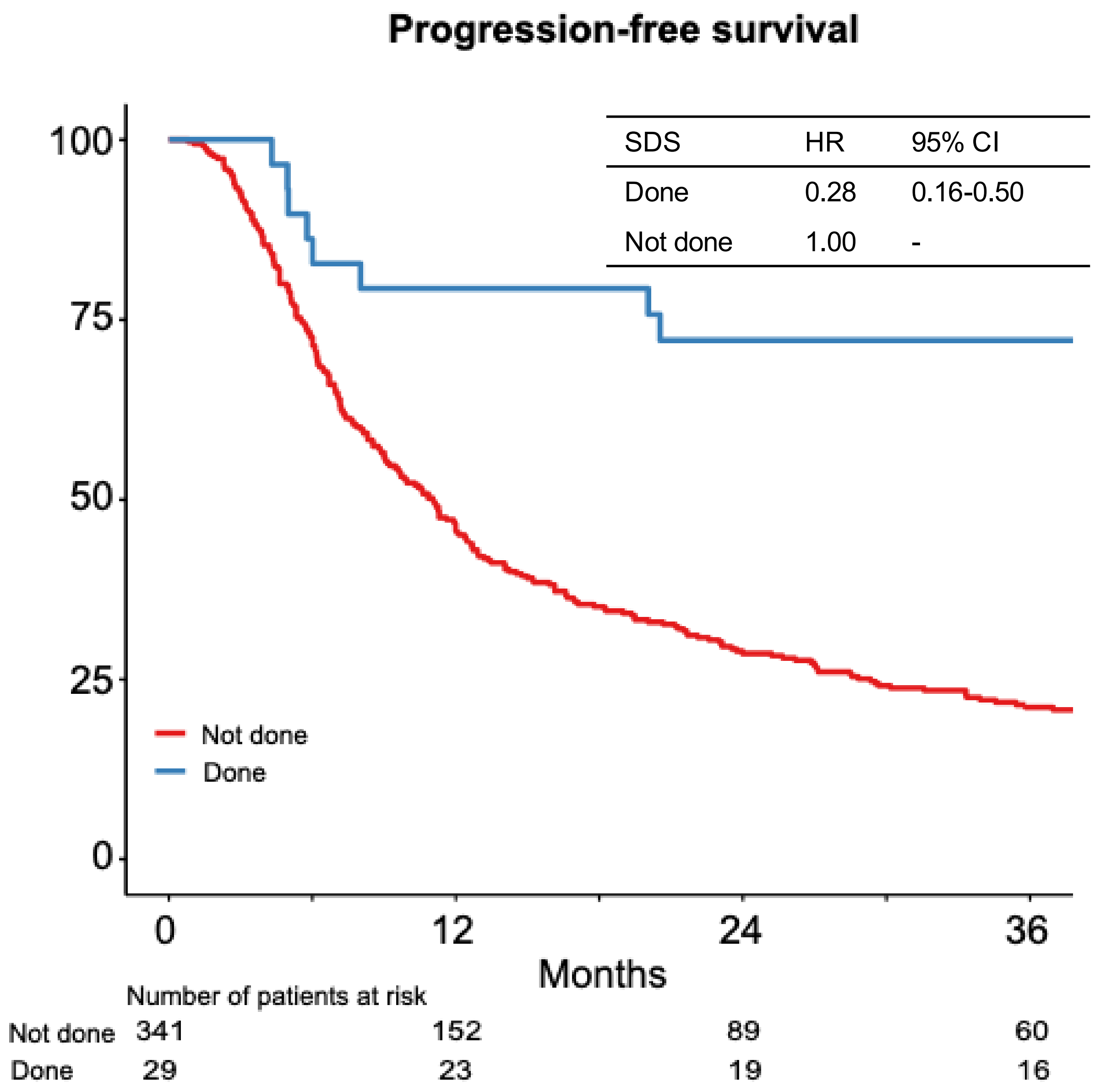
