## Supplementary material for "Real-world safety and effectiveness of olaparib maintenance after first-relapse platinum-sensitive ovarian cancer in Japan: a multicenter historical cohort (JGOG3026)": Table S1

**Supplementary Table 1. A list of 43 JGOG institutions that participated in the JGOG3026 study**

| Institution | N |
| --- | --- |
| Cancer Institute Hospital | 50 |
| Tokyo Metropolitan Cancer and Infectious Diseases Center Komagome Hospital | 28 |
| Kurume University School of Medicine | 22 |
| Niigata University Graduate School of Medical and Dental Sciences | 19 |
| Shizuoka Cancer Center | 19 |
| Faculty of Medicine, University of Tsukuba | 19 |
| Kansai Rosai Hospital | 18 |
| Graduate School of Medicine, Tohoku University | 17 |
| Gifu University Hospital | 16 |
| Kobe City Medical Center General Hospital | 15 |
| University of Occupational and Environmental Health | 14 |
| The Jikei University Katsushika Medical Center | 14 |
| The University of Osaka Hospital | 14 |
| Gunma University Hospital | 13 |
| Kyoto University Hospital | 12 |
| Kitasato University Hospital | 11 |
| Jichi Medical University Hospital | 11 |
| The Jikei University Kashiwa Hospital | 11 |
| Tottori University Hospital | 11 |
| Fukushima Medical University Hospital | 11 |
| Ehime University Hospital | 10 |
| Keio University Hospital | 10 |
| The Jikei University Hospital | 10 |
| Niigata Cancer Center Hospital | 9 |
| Kindai University Hospital | 8 |
| Nara Medical University Hospital | 8 |
| Tokyo Women's Medical University Medical Center East | 7 |
| National Hospital Organization Shikoku Cancer Center | 6 |
| National Cancer Center Hospital East | 6 |
| Tokyo Metropolitan Tama Medical Center | 5 |
| Hiroaki University Hospital | 5 |
| Nagoya University Hospital | 5 |
| Suita Tokushukai Hospital | 4 |
| Hokkaido University Hospital | 4 |
| Hiroshima Prefectural Hospital | 4 |
| Kure Medical Center and Chugoku Cancer Center | 3 |
| Saitama Hospital | 3 |
| Tokai University Hospital | 3 |
| Tokushima University Hospital | 3 |
| Saga University Hospital | 3 |
| The Jikei University Daisan Hospital | 3 |
| Tottori City Hosptal | 2 |
