## Supplementary material for "Real-world safety and effectiveness of olaparib maintenance after first-relapse platinum-sensitive ovarian cancer in Japan: a multicenter historical cohort (JGOG3026)": Table S2

**Supplementary Table 2. Dose modifications and discontinuations owing to adverse events**

|  | mITT1  (n = 413) | mITT2  (n = 284) |
| --- | --- | --- |
| Dose Intensity (mg/day) | 507.5 ± 108.9 | 506.0 ± 113.7 |
| Dose interruption | 161 (39%) | 109 (38%) |
| Dose reductions | 54 (13%) | 34 (12%) |
