## Supplementary material for "Real-world safety and effectiveness of olaparib maintenance after first-relapse platinum-sensitive ovarian cancer in Japan: a multicenter historical cohort (JGOG3026)": Table S3

**Supplementary Table 3. The association between stratified factors and overall survival**

|  | N | Hazard ratio | 95%CI |
| --- | --- | --- | --- |
| Chemotherapy response  CR  PR | 117  293 | 0.73  1.00 | 0.54-0.98  - |
| Germline BRCA pathogenic variants  Yes  No  Unknown | 21  23  368 | 0.30  1.00  1.02 | 0.11-0.83  -  0.60-1.76 |
| Histology  High-grade serous  Others | 322  90 | 0.70  1.00 | 0.52-0.95  - |
| Stage  I-II  III  IV | 60  247  102 | 1.05  1.24  1.00 | 0.67-1.65  0.91-1.71  - |
| Secondary debulking surgery  Done  Not done | 29  345 | 0.26  1.00 | 0.11-0.58  - |
