## Supplementary material for "Real-world safety and effectiveness of olaparib maintenance after first-relapse platinum-sensitive ovarian cancer in Japan: a multicenter historical cohort (JGOG3026)": Table S4

**Supplementary Table 4. Response rate of third-line chemotherapy for recurrent ovarian cancer after olaparib maintenance therapy**

|  |  | CR | | PR | | CR + PR | |
| --- | --- | --- | --- | --- | --- | --- | --- |
| Platinum agent |  | N | RR (95%CI) | N | RR (95%CI) | N | RR (95%CI) |
| Yes | 78 | 3 | 3.8 (0.8-10.8) | 12 | 15.4 (8.2-25.3) | 15 | 19.2 (11.2-29.7) |
| No | 162 | 9 | 5.6 (2.6-10.3) | 26 | 16.0 (10.8-22.6) | 35 | 21.6 (15.5-28.7) |
